# Serum *Mycoplasma pneumoniae* IgG in COVID-19: A Protective Factor

**DOI:** 10.1101/2020.04.12.20060079

**Authors:** Bobin Mi, Lang Chen, Adriana C. Panayi, Yuan Xiong, Guohui Liu

**Affiliations:** Department of Orthopedics, Union Hospital, Tongji Medical College, Huazhong University of Science and Technology, Wuhan 430022, China; Division of Plastic Surgery, Department of Surgery, Brigham and Women’s Hospital and Harvard Medical School, Boston, MA, 02115, USA

## Abstract

**Background:** A correlation between prior exposure to Mycoplasma pneumoniae (IgG positive) and better clinical response to COVID-19 was elusive.

**Methods:** A retrospective review of all COVID-19 infected patients treated at Wuhan Union Hospital from Feb 1 to Mar 20 was carried out. Continuous variables were described as mean, median, and interquartile range (IQR), while categorical variables were compared by X2 test or Fisher’s exact test between COVID-19 infected patients with mycoplasma lgG (-) and mycoplasma lgG (+).

**Results:** Statistically significant differences were shown in terms of laboratory test results. COVID-19 infected patients with mycoplasma lgG positivity had a higher lymphocyte count and percentage (p=0.026, p=0.017), monocyte count and percentage (p=0.028, p=0.006) and eosinophil count and percentage (p=0.039, p=0.007), and a lower neutrophil count and percentage (p=0.044, p=0.006) than COVID-19 infected patients without mycoplasma lgG. Other routine blood tests, including coagulation tests, blood biochemistry and infection-related biomarkers did not significantly differ except for thrombin time (p=0.001) and lactate dehydrogenase (p=0.008). Furthermore, requirement and use of a nasal catheter or oxygen mask was significantly lower in COVID-19 infected patients with mycoplasma lgG positivity (p=0.029).

**Conclusions:** Our findings indicate that mycoplasma IgG positivity is a potential protective factor for SARS-CoV-2 infection.

## Introduction

Ever since its initial outbreak in Wuhan, Hubei province, China, in December 2019, the 2019 novel coronavirus disease (COVID-19) has quickly spread around the world. Currently, more than 1.3 million cases have been confirmed, with 76,420 deaths worldwide.^1^ The clinical characteristics of patients with COVID-19 have been well described with some risk factors shown to increase the mortality of COVID-19 identified, including diabetes, cancer and aging.^2,3^ Protective factors that may help patients with COVID-19 show fewer severe symptoms and better recovery remain elusive. As frontline medical personnel in the Wuhan Union Hospital—one of the biggest COVID-19 designated institutions in Wuhan—we have unique first hand experience in dealing with this pandemic. Here, we report a correlation between prior exposure to *Mycoplasma pneumoniae* (IgG positivity) and better clinical response to COVID-19. This association has, to the best of our knowledge, not been previously shown.

## Methods

A retrospective review of all COVID-19 infected patients treated at Wuhan Union Hospital from Feb 1 to Mar 20 was carried out. COVID-19 was diagnosed in accordance with the New Coronavirus Pneumonia Prevention and Control Program, 7th edition, published by the National Health Commission of China. Clinical symptoms, medical records as well as laboratory tests were reviewed. Continuous variables were described as mean, median, and interquartile range (IQR), while categorical variables were compared by X^2^ test or Fisher’s exact test between COVID-19 infected patients with mycoplasma lgG (-) and mycoplasma lgG (+). Data were expressed using frequency rates and percentages. The Kolmogorov-Smirnov test was used to establish whether continuous variables were normally distributed. Then independent group t test was used if normally distributed, otherwise, the Mann-Whitney test was used. A two-sided α of less than 0.05 was considered statistically significant. SPSS version 23.0 was used for all statistical analyses. This retrospective study was approved by The Institutional Review Board at Union Hospital, Tongji Medical College, Huazhong University of Science and Technology.

## Results

A total of 133 patients with COVID-19 were identified, of which 38 displayed Mycoplasma IgG positivity. Comparison of COVID-19 patients with mycoplasma lgG positivity against those without mycoplasma IgG, showed no differences in the demographics (age, p=.105; female sex, p=.375) or common signs and symptoms of the patients (fever, p=0.23; cough, p=0.53; fatigue, p=1.000; dyspnea, p=0.67; diarrhea, p=0.39; Table 1). Statistically significant differences were, however, shown in terms of laboratory test results. COVID-19 infected patients with mycoplasma lgG positivity had a higher lymphocyte count and percentage (p=0.026, p=0.017), monocyte count and percentage (p=0.028, p=0.006) and eosinophil count and percentage (p=0.039, p=0.007), and a lower neutrophil count and percentage (p=0.044, p=0.006) than COVID-19 infected patients without mycoplasma lgG. Other routine blood tests, including coagulation tests, blood biochemistry and infection-related biomarkers did not significantly differ except for thrombin time (p=0.001) and lactate dehydrogenase (p=0.008). Furthermore, requirement and use of a nasal catheter or oxygen mask was significantly lower in COVID-19 infected patients with mycoplasma lgG positivity (p=0.029), suggesting a better prognosis. The rate of noninvasive ventilation and mortality did not differ; this may be due to lack of relevant clinical cases.

**Table 1.** Demographic characteristics, laboratory findings, treatment and outcomes of COVID-19 infected patients categorized by mycoplasma lgG antibody presence.

|  | Mycoplasma IgG (-)<br>(n=95) | Mycoplasma IgG (+)<br>(n=38) | P value |
| --- | --- | --- | --- |
| **Demographics** |  |  |  |
| Age, y | 66.0 (56.0-70.0) | 61.0 (43.5-70.0) | 0.105 |
| Female sex | 43 (45.3%) | 14 (36.8%) | 0.375 |
| **Signs and symptoms** |  |  |  |
| Fever | 80 (84.2%) | 35 (92.1%) | 0.229 |
| Cough | 70 (73.7%) | 30 (78.9%) | 0.526 |
| Fatigue | 60 (63.2%) | 24 (63.2%) | 1.000 |
| Dyspnea | 18 (18.9%) | 6 (15.8%) | 0.669 |
| Diarrhea | 12 (12.6%) | 7 (18.4%) | 0.389 |
| **Laboratory test results** |  |  |  |
| White blood cell count, x10<sup>9</sup>/L | 5.84 (4.38-7.58) | 5.61 (3.73-7.07) | 0.191 |
| Hemoglobin, g/L | 128.00 (116.00-135.00) | 123.00 (113.50-134.25) | 0.417 |
| Neutrophils, % | 73.40±14.17 | 66.05±12.28 | 0.006* |
| Lymphocytes, % | 16.50 (9.20-26.35) | 21.55 (16.10-30.80) | 0.017* |
| Monocytes, % | 6.83±3.26 | 8.59±3.40 | 0.006* |
| Eosinophils, % | 0.30 (0.10-1.00) | 1.04 (0.20-1.68) | 0.007* |
| Neutrophil count, x10<sup>9</sup>/L | 3.95 (2.92-6.47) | 3.23 (2.40-4.95) | 0.044* |
| Lymphocyte count, x10<sup>9</sup>/L | 0.86 (0.57-1.29) | 1.09 (0.73-1.55) | 0.026* |
| Monocyte count, x10<sup>9</sup>/L | 0.33 (0.25-0.55) | 0.53 (0.30-0.64) | 0.028* |
| Eosinophils count, x10<sup>9</sup>/L | 0.02 (0.00-0.075) | 0.05 (0.30-0.64) | 0.039* |
| D-dimer, mg/L | 0.67 (0.30-2.41) | 0.58 (0.32-1.41) | 0.279 |
| PT, s | 13.40 (12.70-14.20) | 12.95 (12.50-13.63) | 0.069 |
| APPT, s | 37.90 (34.20-42.60) | 35.7 (33.25-40.55) | 0.104 |
| TT, s | 16.03±1.32 | 16.96±1.52 | 0.001* |
| K, mmol/L | 3.83±0.56 | 3.81±0.53 | 0.824 |
| Na, mmol/L | 138.30 (135.75-140.40) | 138.60 (135.35-142.25) | 0.430 |
| ALT, U/L | 33.00 (19.50-55.50) | 31.00 (20.00-46.25) | 0.784 |
| AST, U/L | 36.00 (24.00-53.50) | 29.00 (20.75-39.50) | 0.053 |
| LDH, U/L | 299.00 (215.00-402.25) | 236.00 (192.25-303.75) | 0.008* |
| Total bilirubin, | 12.10 (8.65-15.80) | 10.90 (9.88-15.83) | 0.945 |
| μmol/L |  |  |  |
| Direct Bilirubin, μmol/L | 3.80 (2.75-5.50) | 3.90 (2.78-5.25) | 0.743 |
| Indirect Bilirubin, μmol/L | 7.70 (5.50-9.80) | 7.40 (6.00-10.50) | 0.757 |
| Total Protein, g/L | 62.79±5.62 | 63.18±7.58 | 0.748 |
| Creatinine, μmol/L | 73.10 (58.25-89.45) | 70.00 (62.50-81.83) | 0.632 |
| Blood Urea Nitrogen, mmol/L | 4.41 (3.71-5.96) | 4.23 (3.50-5.04) | 0.181 |
| Ferritin, ng/ml | 644.99 (334.94-1318.25) | 307.16 (202.51-706.99) | 0.136 |
| **Treatment and outcomes** |  |  |  |
| Nasal catheter or oxygen mask | 62 (65.3%) | 17 (44.7%) | 0.029* |
| NIV | 6 (6.3%) | 1 (2.6%) | 0.667 |
| Death | 5 (5.3%) | 1 (2.6%) | 0.843 |

NIV = noninvasive ventilation; Data are presented as n/N (%) or median (IQR). P values were calculated using independent group t test, X<sup>2</sup> test, Fisher's exact test or Mann-Whitney U test. \*p<0.05.
Normal ranges for the laboratory values are as follows: white blood-cell count, 3.5-9.5x10<sup>9</sup>/L; hemoglobin, 130-175 g/L; neutrophil %, 40-75; lymphocyte %, 20-50; monocyte %, 3-10; eosinophils %, 0.4-8; neutrophil count, 1.8-6.3x10<sup>9</sup>/L; lymphocyte count, 1.1-3.2x10<sup>9</sup>/L; monocyte count, 0.1-0.6x10<sup>9</sup>/L; eosinophils count, 0.02-0.52 x10<sup>9</sup>/L; D-dimer, <0.5 ug/ml; PT, 11.0-16.0 s; APTT, 27.0-45.0 s; TT, 14.0-20.0 s; K, 3.5-5.3 mmol/L; Na, 137-147 mmol/L; alanine aminotransferase (ALT), 5-40 U/L; aspartate aminotransferase (AST), 8-40 U/L; lactate dehydrogenase (LDH), 109-245 U/L; total bilirubin, 3.0-20.0 μmol/L; direct bilirubin, 1.7-6.8μmol/L; Total Protein, 60-83g/L; creatinine, 57.0-111.0 μmol/L; Blood Urea Nitrogen, 2.9-8.2mmol/L; ferritin, 21.81-274.66 ng/ml.

## Discussion

Patients with prior exposure to *Mycoplasma pneumoniae* (IgG positive) showed stronger resistance and better recovery from the SARS-CoV-2 viral infection than patients without exposure. Of particular note, the immune response rendered against the virus was noted to be stronger in the IgG positive group, evident through an overall higher leukocyte count. This can partially be explained by reprogrammed immune cells.^4^ Prior evidence has supported that when the lungs recover from infection they develop new alveolar macrophage biology, which can protect the lungs of adults against pneumonia.^5^ Our retrospective patient review highlighted a similar phenomenon: COVID-19 patients with IgG positivity displayed milder symptoms and rarely experienced severe disease (OR, 2.31; 95% CI, 1.89-3.02). To eliminate the interference of age, we compared the outcomes of elder patients (> 60 years) and younger patients (< 30 years) with IgG positivity and found them to be similar (OR, 2.31; 95% CI, 1.89-3.02). Thus, our findings indicate that mycoplasma IgG positivity is a potential protective factor for SARS-CoV-2 infection.

Although these promising observations require further investigation they demonstrate that previous *Mycoplasma pneumoniae* infection is an important part of a patient’s medical history as patients without prior infection are more vulnerable to COVID-19.

## Data Availability

The corresponding authors retain the right to decide whether to share the data based on the research objectives and plan provided by the requesting individual.

## Conflict of Interest Disclosures

None reported.

## Notes

### Competing Interest Statement

The authors have declared no competing interest.

### Funding Statement

Not applicable

## Reference

1. The Center for Systems Science and Engineering at Johns Hopkins University. https://www.arcgis.com/apps/opsdashboard/index.html#/bda7594740fd40299423467b48e9ecf6. Retrieved 04.07.2020.

2. Chen N, Zhou M, Dong X, et al. Epidemiological and clinical characteristics of 99 cases of 2019 novel coronavirus pneumonia in Wuhan, China: a descriptive study. Lancet. 2020;395(10223):507–513.

3. Liang W, Guan W, Chen R, et al. Cancer patients in SARS-CoV-2 infection: a nationwide analysis in China. Lancet Oncol. 2020;21(3):335–337.

4. Jokinen C, Heiskanen L, Juvonen H, et al. Microbial etiology of community-acquired pneumonia in the adult population of 4 municipalities in eastern Finland. Clin Infect Dis. 2001;32(8):1141–1154.

5. Bhagwat SP, Gigliotti F, Wang J, et al. Intrinsic Programming of Alveolar Macrophages for Protective Antifungal Innate Immunity Against Pneumocystis Infection. Front Immunol. 2018;9:2131.

